# An Explainable Analytics Framework for Identifying Process Deviations in Emergency Care for Patients with Multiple Long-Term Conditions

**DOI:** 10.64898/2026.11.24.26368861

**Authors:** Yetong Ding, Yinhuan Dong, Atul Anand, Nazir Lone, Jacques Fleuriot, Tim Kerby, Sean Smith, Ian Underwood

## Abstract

Patients with Multiple Long-Term Conditions (MLTCs), also known as multimorbidity, often present clinical and operational challenges in emergency departments (EDs), yet how their care pathways deviate from expected workflows remains underexplored. This study introduces a process-aware analytical framework that quantifies and interprets ED process deviations by integrating multi-perspective conformance checking with explainable machine learning. Using routinely collected Scottish ED data between 2017–2023, we derived an empirical benchmark pathway. We introduced a new metric, the deviation score (DS), which captures both structural misalignments and breaches of the four-hour ED standard, a national service target defined as completion of ED care within four hours of arrival. Compared with non-MLTC attendances, MLTC patients exhibited longer and more complex pathways, higher DS values, and lower conformance to the benchmark model. The framework enabled interpretable SHAP analyses, revealing that deviation severity among MLTC attendances was primarily associated with triage acuity, year of attendance, activity queue length (number of patients queuing for the same care activity), and referral source, with additional contributions from age, arrival time, and number of chronic conditions. Threshold effects were observed in both queue length and multimorbidity burden, beyond which deviation severity increased sharply, suggesting points where standard pathways become harder to maintain. These findings show that deviation severity for MLTC patients is shaped by clinical urgency and system-level pressures at presentation. However, the inherent complexity of MLTC patients may further increase vulnerability to deviation. The framework offers the potential foundation for process evaluation and targeted improvement in emergency care.

## I. Introduction

**M**ultiple long-term conditions (MLTCs), also known as multimorbidity, refers to the coexistence of two or more chronic health conditions in the same individual [1]. It is estimated that approximately one in three people globally live with multimorbidity [2] [3], [4]. Prevalence increases significantly with age, from just 0.9% in those aged 0–19 to 68.2% among individuals aged 80 and above [5]. People with MLTC are considered clinically complex due to the need for simultaneous management of chronic illnesses that may interact in unpredictable and compounding ways. These challenges are often exacerbated by polypharmacy, socioeconomic vulnerability, and the high burden of self-management [6], [7], [8]. This multifaceted complexity puts pressure on care systems that are traditionally structured around single-disease protocols and episodic treatment models. [6].

Emergency departments (EDs), primarily designed for acute, emergency care [1], [3], often struggle to accommodate the complex needs of MLTC patients [9], [10], [4]. Across the United Kingdom, ED performance is monitored using an access standard, a national time-based target for patients to complete ED care within four hours. This standard influences how patient flow is managed through the system. The tension between this time-driven performance target and the complex clinical needs of patients with MLTC has the potential to distort ED care and result in fragmented delivery [9]. Transitions of care, such as movements between locations or providers, are often slower, less consistent, and more prone to failure [10]. The associated operational challenges may result in process deviations, defined as discrepancies between the expected or recommended pathway based on clinical guidelines or protocols and actual care [8]. Such deviations may involve skipped steps due to time pressures or perceived redundancy, or additional steps introduced in response to complexity or procedural gaps [11]. Whereas some deviations are adaptive and patient-centred [12], [13], others introduce potentially harmful variation that undermines care quality, increases inefficiency, and elevates the risk of adverse outcomes [14], [8], [15]. A growing body of research highlights that process deviations may accumulate without detection, leading to unresolved clinical problems [16] and increased likelihood of repeat ED attendance [17].

Despite its importance, most prior studies have examined deviation indirectly through its consequences, such as delays, overcrowding or long stays [12], rather than by analysing how the care pathway itself diverges from expectations. To study deviation at the level of the process, researchers have increasingly turned to process mining. Process mining reconstructs real-world care pathways from routine data and this approach has been applied to investigate the timeliness of care, inefficiencies in patient flow, and key bottlenecks in ED processes [18] [19] [20]. Such process-mining analyses demonstrate the value of identifying where deviations and delays arise within the ED process. However, process mining alone cannot explain why certain patients are more likely to experience deviation from expected pathways.

Explainable machine learning improves transparency by enabling interpretation of predictive models. A widely used post-hoc interpretability method is SHapley Additive exPlanations (SHAP), which quantifies feature contributions and helps identify clinical or operational drivers of prolonged stays [21], admission risk [22], and extended length of stay [23]. Whereas explainable machine learning can identify factors associated with deviation risk, it does not capture the underlying structure or timing of processes and therefore cannot detect divergence at the process level.

To address the limits of process mining in identifying the clinical and operational factors associated with deviation, and the limits of SHAP in quantifying divergence at the process level, we introduce a new process-aware framework for detecting, measuring, and interpreting the magnitude of process deviations in ED care for patients with MLTC, as illustrated in Fig. 1. We summarise the major contributions of this paper as the following:

- The proposed framework integrates multi-perspective conformance checking with explainable machine learning, enabling both process-level and feature-level understanding of deviation.
- A new deviation metric, Deviation Score (DS). DS is derived through multi-perspective conformance checking, capturing both control-flow misalignments and breaches of the four-hour ED turnaround performance standard.
- The novel results of a systematic analysis of ED process deviation among patients with MLTC. Our work quantifies how far an attendance diverges from the expected care pathway and explains why deviation happens by identifying the clinical, operational, and temporal, factors associated with deviation.
- Evidence that the deviation experienced by MLTC patients follows systematic and clinically plausible patterns. Our work offers new insights into how MLTC interacts with clinical urgency and system-level pressures to influence variability in care.

**Fig. 1.**
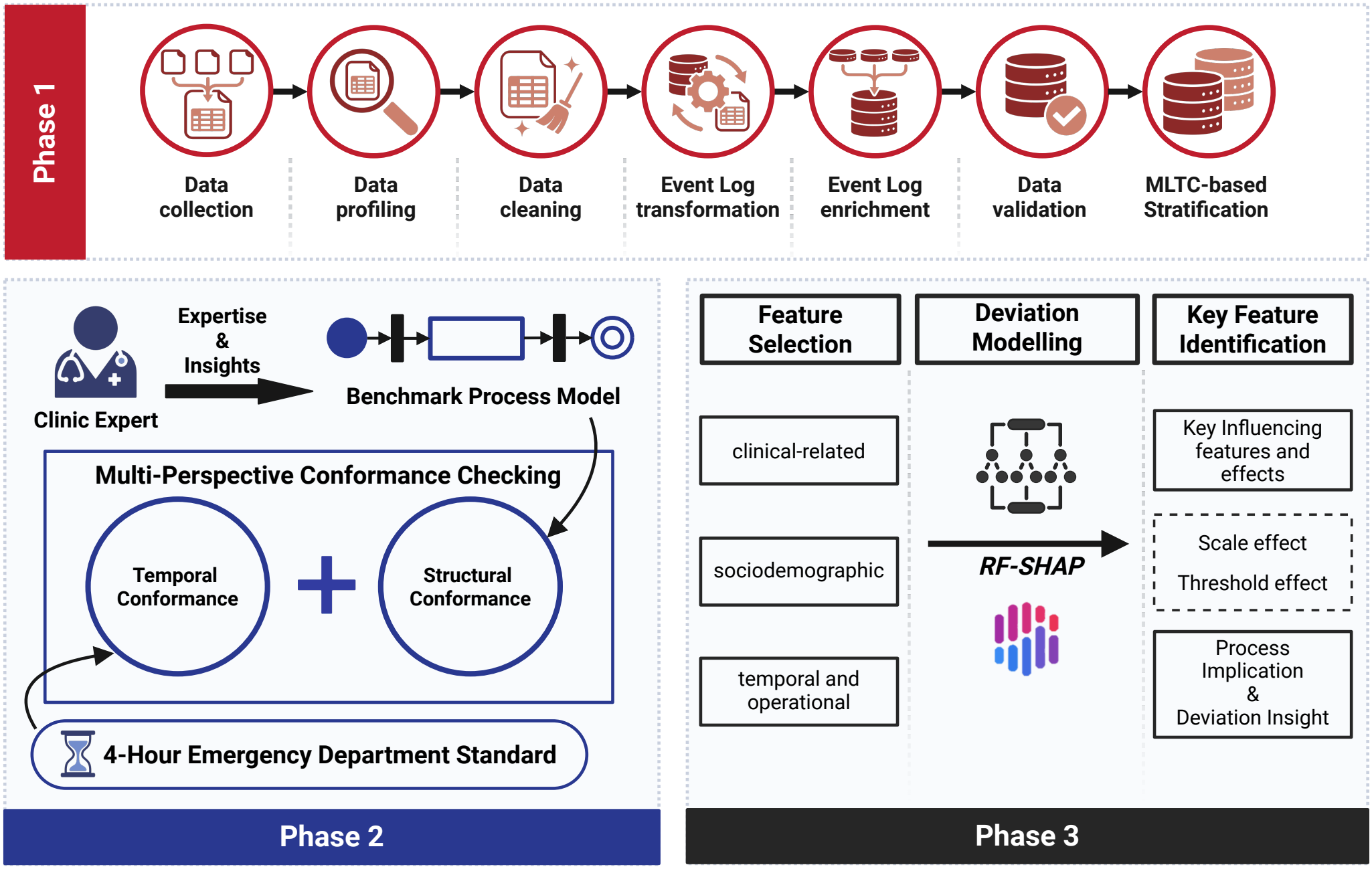
Schematic overview of the proposed process-aware framework for quantifying and explaining ED process deviation for patients with MLTC. Phase 1 (red): data acquisition and preparation. Phase 2 (blue): multi-perspective conformance checking to derive the Deviation Score, capturing both structural and temporal divergence from the expected care pathway. Phase 3 (black): deviation modelling applying explainable machine learning (SHAP) to identify the key clinical and operational drivers that shape deviation severity. Box styles: bold for main stages, solid for feature groups or outputs, and dashed for derived effects.

## II. Related Work

### A. Multiple Long-Term Conditions in Emergency Care

Recent large-scale cohort studies have quantified the burden of MLTC in ED settings. McParland et al. [24] demonstrated that MLTC is linked to both increased ED use and higher mortality. Blayney et al. [12] found a strong association between MLTC and a range of adverse outcomes, including 30-day mortality, re-attendance, and prolonged ED stays. Yordanov et al. [25] argued that ED systems require significant adaptation to meet the needs of MLTC patients, emphasising the importance of pathway-level understanding. Wartelle et al. [26] used cluster analysis to identify distinct multimorbidity profiles among frequent ED users, revealing heterogeneous patterns of service utilization that have implications for targeted interventions. Other work has examined contextual factors influencing ED use, including social determinants [27] and area-level deprivation [28], highlighting inequities in access and outcomes.

Whereas these studies advance understanding of the demand, outcomes, and systemic challenges posed by managing patients with MLTC in EDs, they mainly focus on high-level utilisation metrics [12], outcome disparities [27], [28], and specific intervention points such as triage or discharge planning [10]. Far fewer studies have examined the sequence, timing, and interactions of care activities for this cohort, and none have systematically identified or interpreted process deviations. Moreover, few studies have explored the underlying factors that contribute to such deviations [29]. The lack of pathway-level and deviation-focused evidence limits understanding of process variation in ED care for patients with MLTC.

### B. Process Mining in Emergency Care

Process mining extracts insights from event logs to help analyse, understand, evaluate, and improve real-world processes [30]. It includes three main paths: process discovery (deriving a process model from logs); conformance checking (comparing actual execution against a reference model to detect deviations); and process enhancement (enriching discovered models with performance, resource, or organisational perspectives). Initially developed for business process optimisation, process mining has gained significant traction in healthcare, where it is used to analyse patient pathways, detect inefficiencies, and support evidence-based redesign [31], [32]. In emergency care, most studies emphasize the discovery of control-flow and performance analysis. For example, Duma et al. [33] applied process mining to reconstruct patient flow through the emergency department, identifying bottlenecks and recurrent patterns that contribute to overcrowding. Rojas et al. [34] applied process mining for performance analysis, identifying recurrent loops that extended the length of stay in the ED. Conformance checking has demonstrated potential value in other areas such as inpatient care pathways [35], [36], yet remains comparatively underutilised in ED research. When applied, it typically examines structural (e.g., control-flow) or temporal deviations in isolation. For instance, Cho et al. [20] proposed a set of performance indicators for assessing emergency room processes using process mining. Erdogan et al. [18] conducted one of the few studies that incorporates multiple perspectives, using a goal-driven approach that combines control-flow alignment with performance metrics such as throughput times to identify process bottlenecks.

So, even though prior studies demonstrate the value of conformance checking from a multi-perspective view, there remains a gap in formally quantifying deviation severity from both the expected care pathway and time-based performance targets in ED settings.

### C. Process Mining with Explainable Machine Learning in Emergency Care

Explainable Machine Learning refers to methods that make the predictions of machine learning models understandable to humans, either by using inherently interpretable models or by applying post-hoc techniques such as Shapley Additive Explanations (SHAP) [37]. In healthcare, explainable ML is increasingly valued for translating model outputs into clinically meaningful insights, enhancing trust and usability among practitioners [38]. Existing literature has explored the intersection of explainable ML and process mining. Nannini et al. [37] synthesised approaches for embedding interpretability across the process mining pipeline. Mehdiyev et al. [39] categorized post-hoc and intrinsic techniques for enhancing transparency in process-based predictive models. Whereas these works establish conceptual pathways for integrating process-level analytics with model interpretability, applications in ED settings remain sparse.

In the ED setting, explainable ML has primarily been applied to outcome prediction tasks, with limited attention to process-level analysis. These applications typically focus on identifying feature contributions to specific outcomes,such as prolonged length of stay or admission risk, without examining the underlying care pathways or structural deviations in clinical workflows. For instance, Sulaiman [23] used rule-based explanations to identify operational bottlenecks associated with prolonged length of stay. Arnaud [22] applied SHAP to highlight the key drivers of predicted care trajectories.

Although such approaches strengthen model transparency at the feature level, they do not incorporate formal process-oriented perspectives.

Our work addresses this gap by integrating explainable ML with multi-perspective conformance checking to provide a unified view of both feature-level and process-level deviations, enabling a more comprehensive understanding of deviation severity in ED care for patients with MLTC.

## III. Methods

### A. Datasets and Study Population

In this study, data were sourced from DataLoch, the Trusted Research Environment for analysis of linked, de-identified health records from approximately 1M residents in South East Scotland. Linked data was provided from electronic health records in both primary (GP) and hospital care, specifically:

- Demographic information including age, sex, ethnicity, and socioeconomic deprivation.
- ED attendance and discharge data including activities such as bed request time.
- Hospital inpatient data to understand who required admission from ED.
- National administrative data from the Scottish Morbidity Record for hospital attendances to understand MLTC status of individual patients using ICD-10 coding applied to a subset of 156 CALIBER phenotype condition codelists [40].
- GP data to understand MLTC using Read codes applied to the same subset of 156 CALIBER phenotype condition codelists [40]. These phenotype definitions and associated code lists were applied without modification.
- Laboratory results including time-stamped data on request time.

We included all emergency department attendances presenting to the Royal Infirmary of Edinburgh during the study period between 1st January 2017 to 24th April 2023. In line with the focus of study on characterising pathway variation, each episode of care was treated as an independent case, even when multiple episodes were attributed to the same individual.

### B. Data Pre-processing

Activities of interest include *attendance, triage, blood test, bed request, discharge*, and *inpatient admission*. Each activity was recorded as a single timestamped event, without information on the staff member, role, or organisational unit responsible for performing it. Attendances with incomplete, inconsistent, or implausible temporal information were excluded, including temporal data quality issues such as admission events recorded before ED arrival, bed requests logged after inpatient admission and episodes with missing discharge timestamps. ED stays extending beyond 48 hours and durations between ED discharge and hospital admission that exceeded a statistically defined outlier threshold (above the 99th percentile) were also excluded.

Health conditions present up to but not including the date of each ED attendance were identified using diagnoses recorded in both primary care and hospital care. At each attendance, MLTC status was assigned by counting the number of unique chronic condition phenotypes recorded before the visit. Attendances were classified as MLTC when two or more chronic condition phenotypes were present, whereas those with a single or no conditions were labelled as non-MLTC.

With the final analysis cohort established, we constructed the event log as follows. We defined the finite set of activity labels as *A* = {*A, T, B, R, D, I*}, corresponding to Attendance (A), Triage (T), Blood Test (B), Bed Request (R), Discharge (D), and Inpatient Admission (I), and let *T* denote the set of timestamps. An *event* is an ordered pair *e* = (*a, t*) ∈ *A* × *T*, where *a* is an activity and *t* is its occurrence time. A *trace* for case *c* is a finite, time-ordered sequence 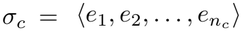, where 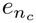 denotes the number of recorded events for case *c*, and with non-decreasing timestamps 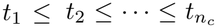. The *event log* is the multiset *L* = {{*σ*_*c*_ | *c* ∈ *C*}}, where *C* denotes the set of all ED attendances included in the study. Each case *c* ∈ *C* is characterised using attributes drawn from three domains: *clinical and care-related, patient sociodemographic*, and *temporal and operational context*. The clinical and care-related domain includes referralSource(*c*), indicating the origin of presentation (e.g., self-referred, GP referral, ambulance). The patient sociodemographic domain includes triageCategory(*c*), a nine-level acuity classification assigned by a triage nurse early in the ED attendance. sex(*c*); ethnicity(*c*); SIMDScore(*c*) derived from the Scottish Index of Multiple Deprivation [41]; nLTC(*c*), the number of Long-Term Conditions (LTCs) identified before the ED visit; and ageGroups(*c*), grouped into ordered age bands (0–17, 18– 30, 31–64, 65–79, 80+). The temporal and operational domain comprises activity-level attributes: arrivalShift(*a*), indicating the shift during which the patient arrived (e.g., day, evening, or night), weekdayWeekend(*a*), season(*a*), year(*a*), and activityQueueLength(*a*), which is calculated as the number of patients concurrently performing activity *a* in the ED. These activity-level features are aggregated to the case level for downstream modelling: the median across activities is used for activityQueueLength(*a*), whereas the first observed value is used for the remaining activity-level attributes.

### C. Benchmark Process Model

The benchmark process model represents the common ED care pathway, defined through empirical frequency analysis of observed event sequences and guided by clinical expertise. The model captures the most common low-complexity trajectory observed in the data, namely *Attendance* → *Triage* → *Discharge*. Activities such as blood tests, bed requests, or inpatient admissions were excluded from the optimised model and, when present, were treated as deviations during conformance analysis.

The benchmark pathway is specified as a Petri net [42] *M* = (*P, T, F, m*_0_, *m*_*f*_ ), shown in Fig. 2, where: *P* = {*p*_0_, *p*_1_, *p*_2_, *p*_3_, *p*_4_, *p*_5_} is the set of *places*, represented graphically as circles, each denoting a state or condition between activities (e.g., after *Attendance* but before *Triage*). *T* = *T*_obs_ ∪ *T*_*τ*_ is the set of *transitions*, where *T*_obs_ = {*A, T, D*} are observable clinical activities (drawn as labelled rectangles) and *T*_*τ*_ = {*τ*_1_, *τ*_2_} are *silent transitions* (drawn as solid black rectangles) introduced to ensure structural soundness and flexible alignment. *F* ⊆ (*P* ×*T* )∪(*T* ×*P* ) is the flow relation. *m*_0_ is the initial marking with *m*_0_(*p*_0_) = 1 and *m*_0_(*p*_*i*_) = 0 for all *i* ≠ 0, representing the entry of a single case trace into the process. *m*_*f*_ is the final marking designating completion of the case when a token (representing the progression of a case through the model) reaches *p*_5_.

**Fig. 2.**
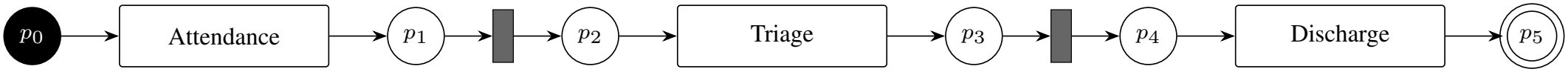
Schematic representation of benchmark normative ED pathway as a Petri net. Key observable clinical activities (▭) follow *Attendance* ***→*** *Triage* ***→*** *Discharge*. Silent transitions ***τ*** (█) ensure structural soundness and alignment flexibility. Intermediate places are shown as (◯), with start marking ***m***_**0**_ and final marking ***m***_***f***_ denoted by (⬤) and (◯), respectively.

### D. Multi-Perspective Conformance Checking

Multi-perspective conformance checking was applied to quantify process deviations by jointly assessing (i) structural adherence to a benchmark ED pathway and (ii) compliance with the ED 4-hour standard. This approach ensured that both control-flow and time-related requirements were incorporated into the analysis.

#### 1) Structural Conformance

For each case trace *c* ∈ *C*, structural conformance is assessed using the alignment-based replay algorithm in PM4Py [43], an open-source Python library for process mining. The algorithm aligns the observed sequence *σ*_*c*_ to the benchmark model *M* by minimizing an *alignment cost*. A uniform unit cost function is adopted, whereby synchronous moves (when an observed event matches a corresponding model transition) incur zero cost, while both log-only moves (events observed in the trace but not enabled in the model) and model-only moves (model transitions not observed in the trace) incur equal unit penalties.

Whereas the alignment cost *C*_align_(*c*) quantifies the absolute degree of deviation for case *c*, it depends on trace length and is therefore not directly comparable across cases. To obtain a normalized and interpretable measure, we compute the standard alignment-based *fitness score* [44], defined as:

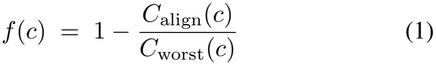

where:

- *C*_align_(*c*) denotes the optimal alignment cost of case *c*, and
- *C*_worst_(*c*) denotes the worst-case alignment cost under the specified cost function, representing the maximum possible deviation for that trace.

The resulting score *f* (*c*) ∈ [0, 1] expresses the degree of structural conformance of case *c* to the benchmark pathway. A value of *f* (*c*) = 1 indicates that the trace *σ*_*c*_ can be replayed on *M* without any deviations, representing perfect conformance, whereas *f* (*c*) = 0 corresponds to the limiting case where every move incurs penalties. Intermediate values *f* (*c*) ∈ (0, 1) capture partial conformance, quantifying the proportion of behaviour in *σ*_*c*_ that is structurally consistent with the normative model. At the cohort level, conformance can also be summarised by the *fitness-based conformance rate*, defined as the proportion of cases with *f* (*c*) = 1, that is, traces that align perfectly with the benchmark pathway.

#### 2) Temporal Conformance

Temporal conformance is assessed relative to the ED 4-hour operational standard, with threshold *T*_max_ = 240 minutes. For each case *c* ∈ *C*, the observed duration is defined as *D*(*c*) = *t*^end^(*c*) − *t*^start^(*c*) where *t*^start^(*c*) corresponds to the timestamp of the *A (Attendance)* event and *t*^end^(*c*) corresponds to the timestamp of the *D (Discharge)* event in the trace *σ*_*c*_. These timestamps represent the recorded start and end of the ED episode and therefore define the interval over which temporal conformance is evaluated.

To quantify deviation beyond the 4-hour standard, we define the temporal penalty TP(*c*) as

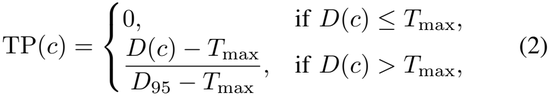

where *D*_95_ denotes the 95th percentile of observed durations across the full study population, serving to normalise extreme values and constrain the penalty within [0, 1]. A value of TP(*c*) = 0 indicates perfect temporal compliance with the NHS 4-hour standard, TP(*c*) = 1 represents the worst observed overruns (beyond the 95th percentile), and intermediate values capture the relative degree of temporal deviation.

#### 3) Multi-Perspective Deviation Metric

To capture deviations that arise either from pathway misalignment or from breaching the 4-hour ED standard, we define the unified deviation metric, Deviation Score (DS), that integrates both structural and temporal perspectives. For each case *c*, the deviation metric is defined as

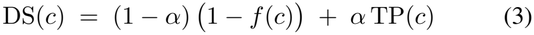

where *f* (*c*) denotes the fitness score, TP(*c*) is the temporal penalty, and *α* ∈ [0, 1] is a weighting parameter that balances the relative contribution of structural versus temporal deviation. We set *α* = 0.5, thereby assigning equal weight to structural and temporal perspectives. This symmetric weighting was adopted as a transparent and neutral baseline in the absence of established theoretical or empirical guidance favouring one dimension of deviation over the other in ED conformance assessment. Similar balanced weighting strategies have been employed in related domains, such as process trace clustering where activity sequences and temporal attributes are jointly considered, symmetric weighting has been reported to yield more stable and interpretable results [45].

The metric is normalised to the interval [0, 1], where DS(*c*) = 0 denotes no deviation in both structural and temporal dimensions, and DS(*c*) = 1 corresponds to the most extreme observed deviation, i.e., maximal misalignment combined with maximum temporal overrun. Intermediate values represent partial deviation, reflecting the relative contribution of structural and temporal departures from the benchmark pathway. For cohort-level reporting, two global measures are used: (i) the *deviation-based conformance rate*, defined as the proportion of cases with DS(*c*) = 0, and (ii) the distribution of DS(*c*), summarised by its median and interquartile range (Q1–Q3).

### E. Feature Selection

To explore which case-level attributes were most strongly associated with variation in the DS, we performed a correlation analysis using Spearman’s rank correlation coefficient along-side Mutual Information. Spearman’s rho captures monotonic relationships between each feature and the continuous DS, whereas Mutual Information identifies non-linear or non-monotonic associations that correlation alone may miss. This exploratory analysis was not used to remove variables. Rather, it provided an initial assessment of which features exhibited meaningful relationships with deviation severity, as well as whether any strong dependencies existed between features.

Because the full feature set comprised only twelve variables across clinical, sociodemographic, and operational domains and each variable carries interpretive relevance for understanding variation in ED pathways, all features were retained for downstream modelling. This approach avoids premature feature exclusion and ensures that the Random Forest models introduced in Section III-F are able to capture vary subtle, context-dependent relationships among features.

To assess multicollinearity among predictors, we also reported Variance Inflation Factors (VIFs) after encoding and scaling all features included in the regression model. This diagnostic step was used to assess whether any variables exhibited strong linear dependence on others. A threshold of VIF *>* 5 was prespecified to indicate strong linear dependence. No variables exceeded this threshold, and therefore no features were excluded.

### F. Deviation Modelling

Deviation severity was modelled as a continuous outcome using a Random Forest (RF) regressor, with the unified DS as the target variable. For each case *c*, the input vector *x*(*c*) comprised the full set of clinical, sociodemographic, and operational attributes described in Section III-B. This modelling framework captures nonlinear relationships and interactions without imposing parametric assumptions about how features relate to deviation severity. Model performance was quantified using Mean Absolute Error (MAE), which reflects the average prediction error on the 0–1 deviation scale, and the coefficient of determination (*R*^2^), which measures the proportion of variance in deviation severity explained by the model. All performance metrics were estimated using 10-fold cross-validation to ensure that MAE and *R*^2^ reflected out-of-sample predictive behaviour.

Because temporal and system-level factors may influence ED workflow dynamics, we further examined the sensitivity of model performance and feature rankings to different representations of the *Year* attribute. Three variants of the RF model were therefore trained: one using year as a numeric variable, one using a clustered encoding distinguishing pre-pandemic (2017-2019), pandemic (2020-2021), and post-pandemic (2021-2023) periods, and one excluding the year variable altogether. This sensitivity analysis allowed us to evaluate how temporal structure contributed to deviation severity and whether alternative encodings of year altered the relative importance of other predictors.

To interpret the RF regression models and ensure transparency, we applied SHAP values computed using the Tree-SHAP algorithm. SHAP provides an additive decomposition of each model prediction into contributions from individual features. For a given case *c* with feature vector *x*(*c*), the model output can be expressed as

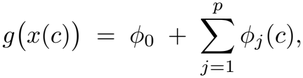

where *g*(·) is the model’s decision function, *ϕ*_0_ is the baseline (expected model output over the training set), and each SHAP value *ϕ*_*j*_(*c*) represents the marginal contribution of feature *j* to the prediction for case *c*, computed as the average effect of adding *j* across all possible subsets of features.

## IV Results

### A. Cohort Description

A total of 427,497 ED attendances were included in the analysis, comprising 315,081 (73.7%) MLTC attendances and 112,416 (26.3%) non-MLTC attendances. Patient characteristics, as defined in Section III-B, were compared between groups and are summarised in Table I. Attendances with MLTC were generally older, more likely to arrive via emergency ambulance and result from primary or speciality care referral (*p <* 0.001). MLTC attendances were also assigned higher-acuity triage categories, whereas non-MLTC attendances were more frequently classified into lower-acuity categories or redirection pathways (*p <* 0.001). Significant cohort differences were additionally observed across sex, SIMD, ethnicity, season, weekday vs weekend, arrival shift, and year of attendance (all *p <* 0.001).

**Table I.** Cohort characteristics of ED attendances by MLTC status. Continuous variables reported as median [IQR]; categorical as ***n*** (%). ***p***-values from Mann–Whitney ***U*** (continuous) and ***χ***^**2**^ (categorical) with FDR adjustment.

| **Variable** | **MLTC** | **Non-MLTC** | *p*-**value** |
|---|---|---|---|
| **Number of attendances** | 315 081 | 112 416 |  |
| **Age Groups – n (%)** |  |  | < 0.001 |
| 18–30 | 34 900 (11.1) | 46 899 (41.7) |  |
| 31–64 | 148 533 (47.1) | 61 150 (54.4) |  |
| 65–79 | 73 789 (23.4) | 3 920 (3.5) |  |
| 80+ | 57 859 (18.4) | 447 (0.4) |  |
| **Sex – n (%)** |  |  | < 0.001 |
| Female | 172 271 (54.7) | 52 656 (46.8) |  |
| Male | 142 810 (45.3) | 59 760 (53.2) |  |
| **SIMD score – n (%)** |  |  | < 0.001 |
| Quintile 1 (most deprived) | 61 933 (19.7) | 19 928 (17.7) |  |
| Quintile 2 | 83 476 (26.5) | 27 084 (24.1) |  |
| Quintile 3 | 50 684 (16.1) | 18 101 (16.1) |  |
| Quintile 4 | 48 782 (15.5) | 19 308 (17.2) |  |
| Quintile 5 (least deprived) | 70 206 (22.3) | 27 995 (24.9) |  |
| **Ethnicity – n (%)** |  |  | < 0.001 |
| White | 296 203 (94.0) | 99 128 (88.2) |  |
| Asian | 5 810 (1.8) | 4 803 (4.3) |  |
| Black | 1 373 (0.4) | 1 380 (1.2) |  |
| Mixed | 1 171 (0.4) | 1 116 (1.0) |  |
| Other | 3 025 (1.0) | 3 288 (2.9) |  |
| Not Stated | 7 499 (2.4) | 2 701 (2.4) |  |
| Number of Long-Term Conditions (LTCs) – median (IQR) | 5 [3–9] | 1 [0–1] | < 0.001 |
| Activity Queue Length – median (IQR) | 3 [1-5] | 2 [1–5] | < 0.001 |
| **Referral source – n (%)** |  |  | < 0.001 |
| Self Referral | 110 320 (35.0) | 71 076 (63.2) |  |
| Emergency Ambulance | 125 074 (39.7) | 15 046 (13.4) |  |
| Primary or speciality care referral | 46 896 (14.9) | 11 215 (10.0) |  |
| NHS24 referral | 20 207 (6.4) | 10 892 (9.7) |  |
| Planned reattendance | 3 466 (1.1) | 1 602 (1.4) |  |
| Other | 9 118 (2.9) | 2 585 (2.3) |  |
| **Triage Category – n (%)** |  |  | < 0.001 |
| 1 – Immediate Resuscitation | 28 102 (8.9) | 2 498 (2.2) |  |
| 2 – Very Urgent | 56 841 (18.0) | 7 467 (6.6) |  |
| 3 – Urgent | 154 122 (48.9) | 45 720 (40.7) |  |
| 4 – Standard | 43 042 (13.7) | 27 252 (24.2) |  |
| 5 – Non Urgent | 696 (0.2) | 429 (0.4) |  |
| 6 – See and Treat | 29 586 (9.4) | 26 635 (23.7) |  |
| 7 – Suitable for redirection | 2 692 (0.9) | 2 415 (2.1) |  |
| **Arrival shift (24h) – n (%)** |  |  | < 0.001 |
| Day (8AM–4PM) | 142 303 (45.2) | 49 984 (44.5) |  |
| Evening (4PM–12AM) | 121 373 (38.5) | 46 078 (41.0) |  |
| Overnight (12AM–8AM) | 51 405 (16.3) | 16 354 (14.5) |  |
| **Year – n (%)** |  |  | < 0.001 |
| 2017 | 49 576 (15.7) | 20 635 (18.4) |  |
| 2018 | 50 091 (15.9) | 18 959 (16.9) |  |
| 2019 | 52 855 (16.8) | 19 556 (17.4) |  |
| 2020 | 48 394 (15.4) | 15 267 (13.6) |  |
| 2021 | 50 043 (15.9) | 16 518 (14.7) |  |
| 2022 | 47 252 (15.0) | 15 966 (14.2) |  |
| 2023 | 16 870 (5.4) | 5 515 (4.9) |  |
| **Season – n (%)** |  |  | < 0.001 |
| Winter | 79 609 (25.3) | 27 858 (24.8) |  |
| Spring | 83 811 (26.6) | 29 734 (26.4) |  |
| Summer | 77 637 (24.6) | 28 242 (25.1) |  |
| Autumn | 74 024 (23.5) | 26 582 (23.6) |  |
| **Weekday vs. Weekend – n (%)** |  |  | < 0.001 |
| Weekday | 227 889 (72.3) | 79 457 (70.7) |  |
| Weekend | 87 192 (27.7) | 32 959 (29.3) |  |

### B. Process and Duration

Differences in ED process pathways between MLTC and non-MLTC attendances were examined using Directly-Follows Graphs (DFGs), which represent observed transitions between consecutive activities, constructed from the 95th percentile of cases to provide a representative view of typical pathways while reducing the influence of rare or atypical cases (Fig. 3). Non-MLTC attendances followed a streamlined pathway, most commonly moving from *Attendance* to *Triage* and then directly to *Discharge*, with infrequent detours through blood testing, bed requests, or inpatient admission. In contrast, MLTC attendances exhibited a markedly more complex and varied workflow, with substantially higher frequencies of transitions involving *Blood Test, Bed Request*, and *Inpatient Admission*, and a more highly branched process structure overall.

**Fig. 3.**
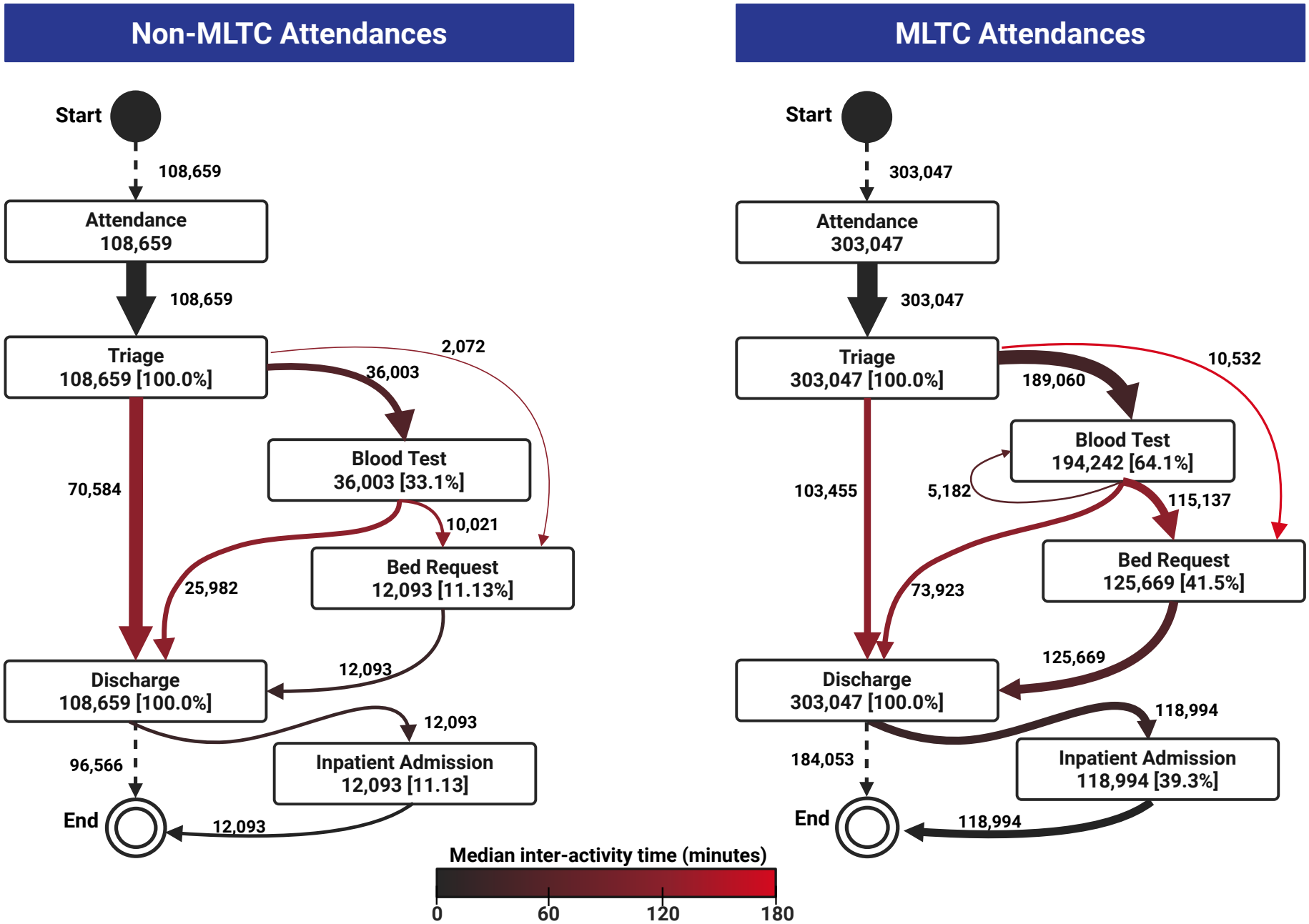
Schematic comparison of ED process models by MLTC status. Arrow thickness denotes frequency of transitions, and colour represents median inter-activity time (minutes). MLTC attendances involve more blood tests, bed requests, and admissions, with longer delays across transitions.

The performance models further indicated that median interactivity times were consistently longer for MLTC attendances, including slower progression from *Triage* to subsequent care events, longer intervals between *Bed Request* and downstream activities. These findings show that MLTC attendances not only traverse a more intricate sequence of ED events but also progress more slowly through key stages of care. As shown in Fig. 4, MLTC attendances experienced longer total ED durations, with a broader spread toward higher values and a greater proportion of cases exceeding the 4-hour operational standard compared with non-MLTC attendances.

**Fig. 4.**
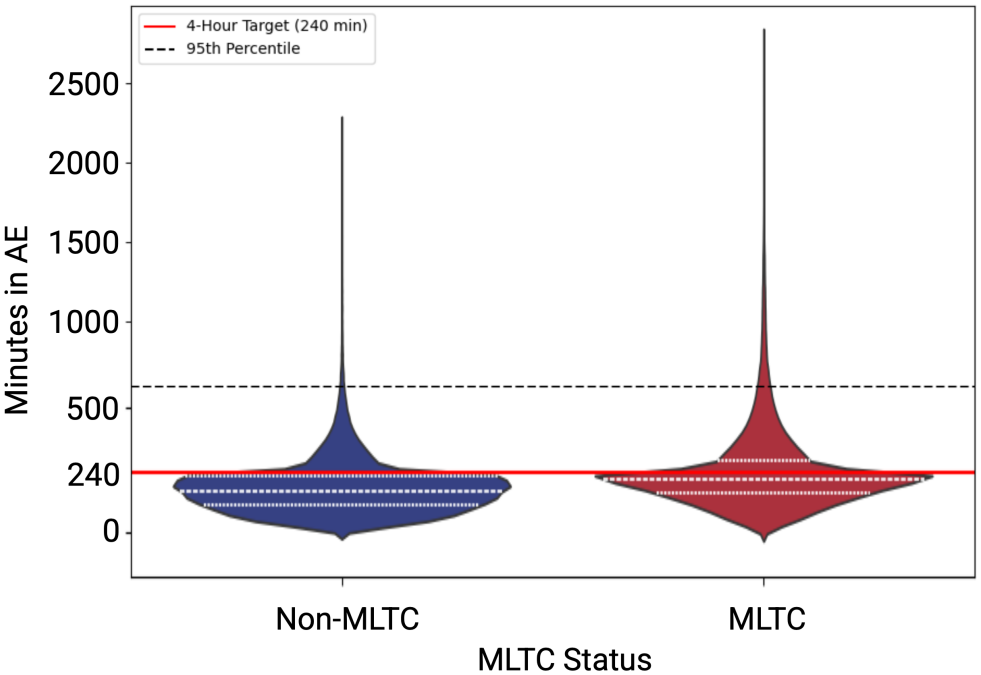
Distribution of ED duration by MLTC status at the case level. The red line marks the NHS 4-hour standard and the black dotted line marks the 95th percentile. The white dashed lines mark the quartiles of the distribution (25th, 50th/median, and 75th percentiles).

### C. Conformance Checking Results

Conformance checking results are summarised in Table II. Conformance rate refers to the proportion of ED attendances that fully aligned with the benchmark pathway, as defined in Section III-D. We report both the fitness-based and deviation-based conformance rates, alongside the distribution of deviation scores. MLTC attendances showed substantially lower conformance than non-MLTC attendances, with fitness-based conformance rates approximately half as high and similarly reduced conformance when assessed using the deviation-based metric. DS were also higher in the MLTC cohort, with a nonzero median and a wider interquartile range, indicating greater variability and overall higher levels of process deviation compared with non-MLTC attendances.

**TABLE II.**
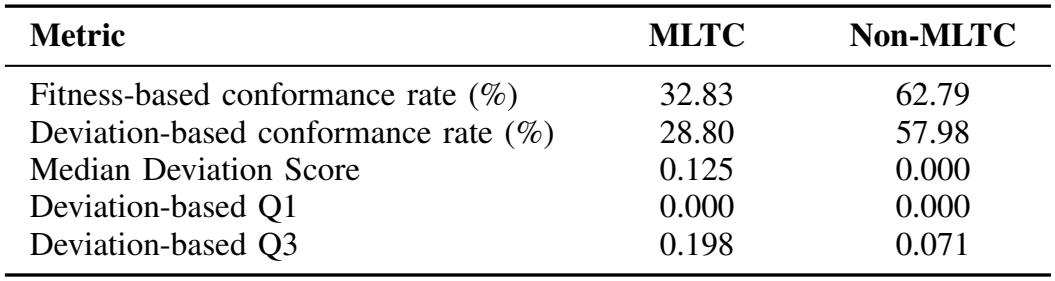
Conformance checking results for MLTC and non-MLTC attendances.

| **Metric** | **MLTC** | **Non-MLTC** |
|---|---|---|
| Fitness-based conformance rate (%) | 32.83 | 62.79 |
| Deviation-based conformance rate (%) | 28.80 | 57.98 |
| Median Deviation Score | 0.125 | 0.000 |
| Deviation-based Q1 | 0.000 | 0.000 |
| Deviation-based Q3 | 0.198 | 0.071 |

### D. Feature Correlation Analysis

To characterise the relationship between each case-level attribute and DS, we computed Spearman’s rank correlation coefficient and Mutual Information (MI) for all features. Results are summarised in Table III. Both measures showed consistent patterns, with the strongest associations observed for *activity queue length (median), triage category, referral source, year pattern*, and *number of LTCs*. Although effect sizes were generally modest, as expected for complex operational outcomes, each of the top predictors exhibited statistically significant relationships with the deviation score. We also examined correlations among features using the correlation matrix. Most feature pairs demonstrated weak or negligible correlation, indicating limited redundancy across clinical, sociodemographic, and operational domains. The strongest correlation was observed between age group and number of LTCs (approximately *ρ* = 0.52), which aligns with clinical expectations that multimorbidity increases with age. Despite this moderate association, all VIF values were close to 1, confirming that no feature showed substantial linear dependence on others and supporting the stability of individual feature contributions.

**TABLE III.** Association between each feature and the deviation score, quantified using Spearman‘s rho and Mutual Information (MI).

| **Feature** | **Spearman's ρ** | **Mutual Information** | **Variance Inflation Factor** |
|---|---|---|---|
| Activity Queue Length (median) **(AQL)** | 0.2500 | 0.4639 | 1.0769 |
| Triage Category **(TC)** | -0.5556 | 0.2902 | 1.3395 |
| Referral Source **(RS)** | -0.3978 | 0.1336 | 1.3245 |
| Year (clustered) **(Y)** | -0.0997 | 0.0883 | 1.0247 |
| Number of LTCs **(NLTC)** | 0.3267 | 0.0668 | 1.4010 |
| Age Group **(AG)** | 0.2789 | 0.0501 | 1.4346 |
| Arrival Shift **(AS)** | 0.1339 | 0.0153 | 1.0616 |
| Season **(SN)** | 0.0051 | 0.0043 | 1.0004 |
| Sex **(SX)** | 0.0142 | 0.0024 | 1.0043 |
| Ethnicity **(E)** | 0.0112 | 0.0007 | 1.0052 |
| SIMD Score **(SIMD)** | 0.0201 | 0.0000 | 1.0635 |
| Weekday vs Weekend **(WW)** | -0.0051 | 0.0000 | 1.0078 |

### E. Regression Performance with Sensitive Analysis

Table IV presents the regression performance of the Random Forest models using three representations of the *year* variable: as a numeric feature, as a clustered categorical variable, and excluded entirely. In the test set, approximately 30% of attendances met the ideal criteria (DS = 0), consistent with the distribution shown in Table II. Across all configurations, MAE values remained low (approximately 0.083–0.093), indicating that the model predicts deviation scores with average errors of less than 0.10 on the 0–1 scale of the target. Corresponding *R*^2^ values ranged from 0.30 to 0.45, showing that a meaningful proportion of the variance in the DS is explained by the clinical, sociodemographic, and operational features included in the model, despite the inherent complexity of ED processes.

**TABLE IV.** Cross-VALIDATED Random Forest regression performance under three representations of the *Year* variable.

| **Model** | **Mean MAE** | **SD MAE** | **Mean** *R*<sup>2</sup> |
|---|---|---|---|
| Year Numeric | 0.0907 | 0.0005 | 0.4452 |
| Year Cluster | 0.0921 | 0.0005 | 0.4336 |
| Year Excluded | 0.1049 | 0.0004 | 0.3122 |

The sensitivity analysis showed that model performance was highly consistent when *year* was encoded either numerically or using clustered temporal groups, with both variants yielding nearly identical MAE and *R*^2^ estimates. In contrast, removing the *year* variable resulted in a noticeable increase in MAE and a substantial reduction in *R*^2^, highlighting that temporal context contributes meaningfully to explaining variation in deviation severity.

### F. SHAP Interpretation of Deviation Modelling

Shapley value analysis was used to interpret the Random Forest regression model and identify the most influential factors driving deviation severity. In this context, positive SHAP values correspond to increased predicted deviation severity, whereas negative values correspond to decreased predicted deviation. As shown in Fig. 5, the features with the greatest average contribution to the model output were *triage category, year (clustered), activity queue length (median), referral source*, and *age group*.

**Fig. 5.**
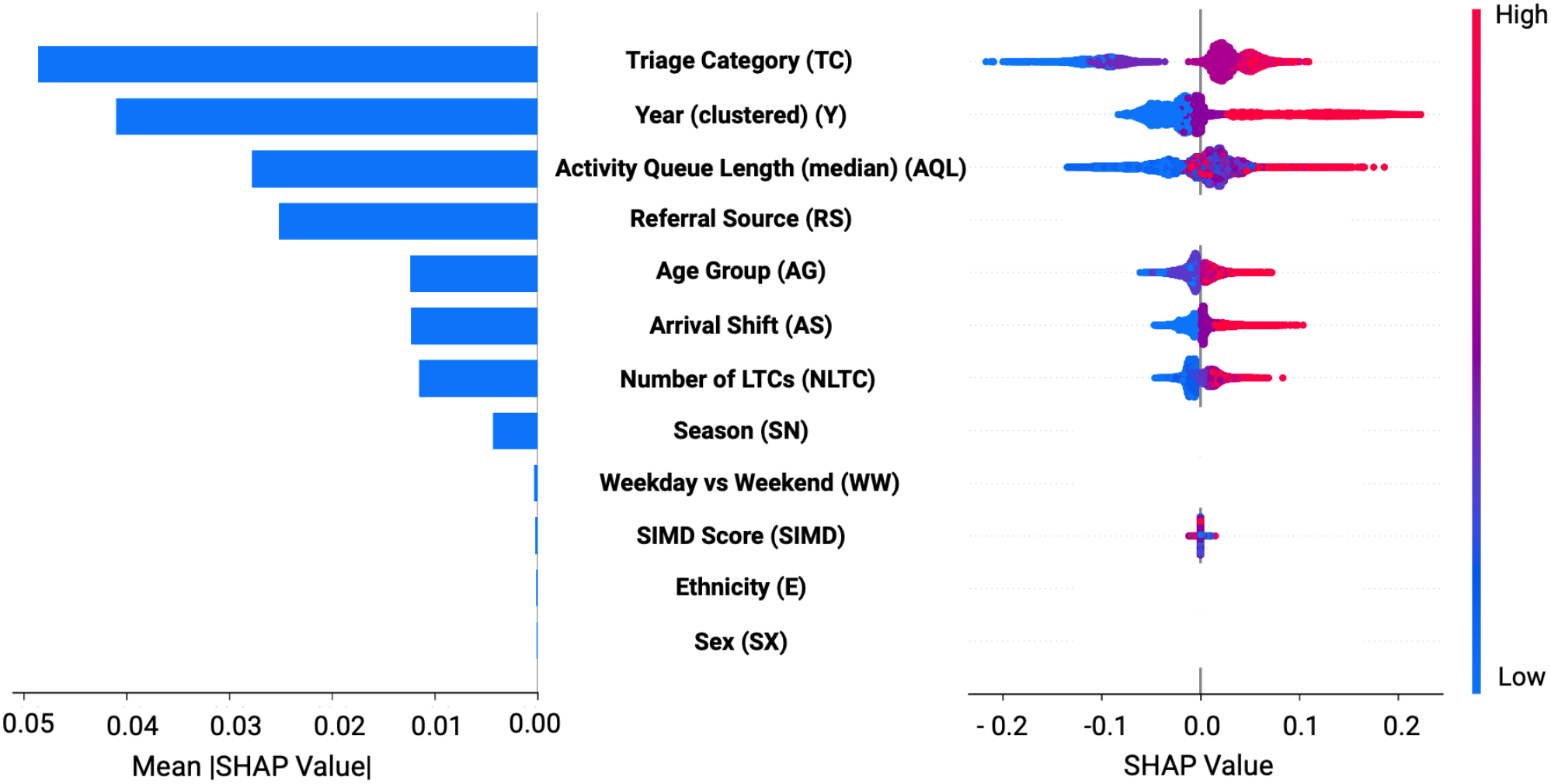
SHAP summary plot showing the distribution and magnitude of Shapley values for each feature.

Triage category (TC) was the dominant predictor, with higher-acuity cases (e.g., Immediate Resuscitation (Code 1)) consistently associated with increased deviation scores. Year (clustered) also showed a clear pattern, where attendances occurring in later pandemic and post-pandemic periods were associated with increased deviation when compared with earlier years. Operational pressure, as captured by median activity queue length at time of attendance, was another key driver, with higher queue lengths consistently associated with higher predicted deviation. Referral source contributed substantially to prediction but, being nominal, does not display a colour gradient in the dot plot; its category-specific effects are shown in Fig. 6. Emergency ambulance and primary or speciality care referral tended to increase deviation, whereas other categories showed predominantly negative SHAP values.

**Fig. 6.**
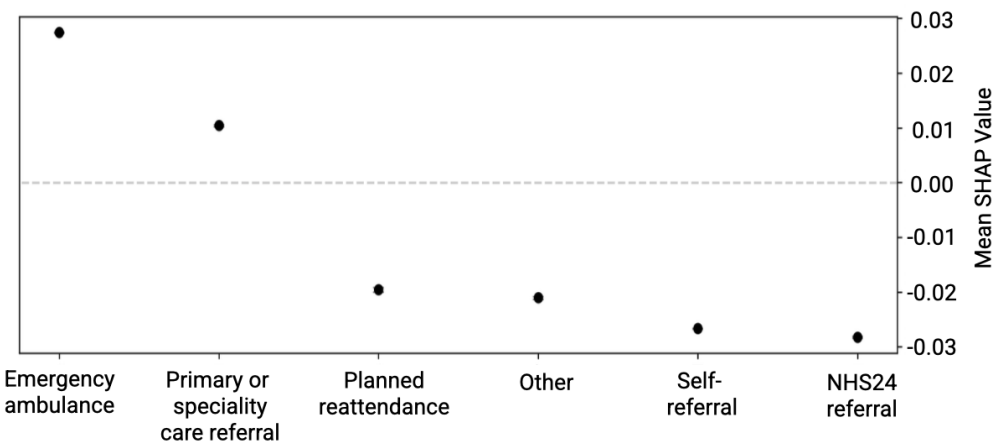
SHAP value distributions for each category of *ReferralSource*.

Age group, arrival shift, and number of LTCs contributed at a similar level to model output. Whereas age group appeared among the top five features in the SHAP summary plot, all three variables demonstrated moderate and consistent influence on deviation severity. Age group effects were generally monotonic, with older patients associated with higher predicted deviation. The distribution of arrival shift suggested that arrival outside of standard hours (evening and overnight) where ED staffing profiles are different, was associated with higher deviation. Although all patients in the cohort had at least two long-term conditions, variation in the number of LTCs still accounted for meaningful differences in predicted deviation. Sociodemographic and temporal factors such as sex, ethnicity, SIMD score, season, and weekday vs weekend had comparatively smaller SHAP values, indicating limited influence on deviation severity relative to operational and clinical predictors.

SHAP dependence plots (Fig. 7) were generated to further interpret the contribution and interactions of key predictors. Each plot represents how changes in a feature’s value influenced its SHAP contribution to the predicted deviation severity, and colour scales indicates the interacting feature with the strongest estimated effect.

**Fig. 7.**
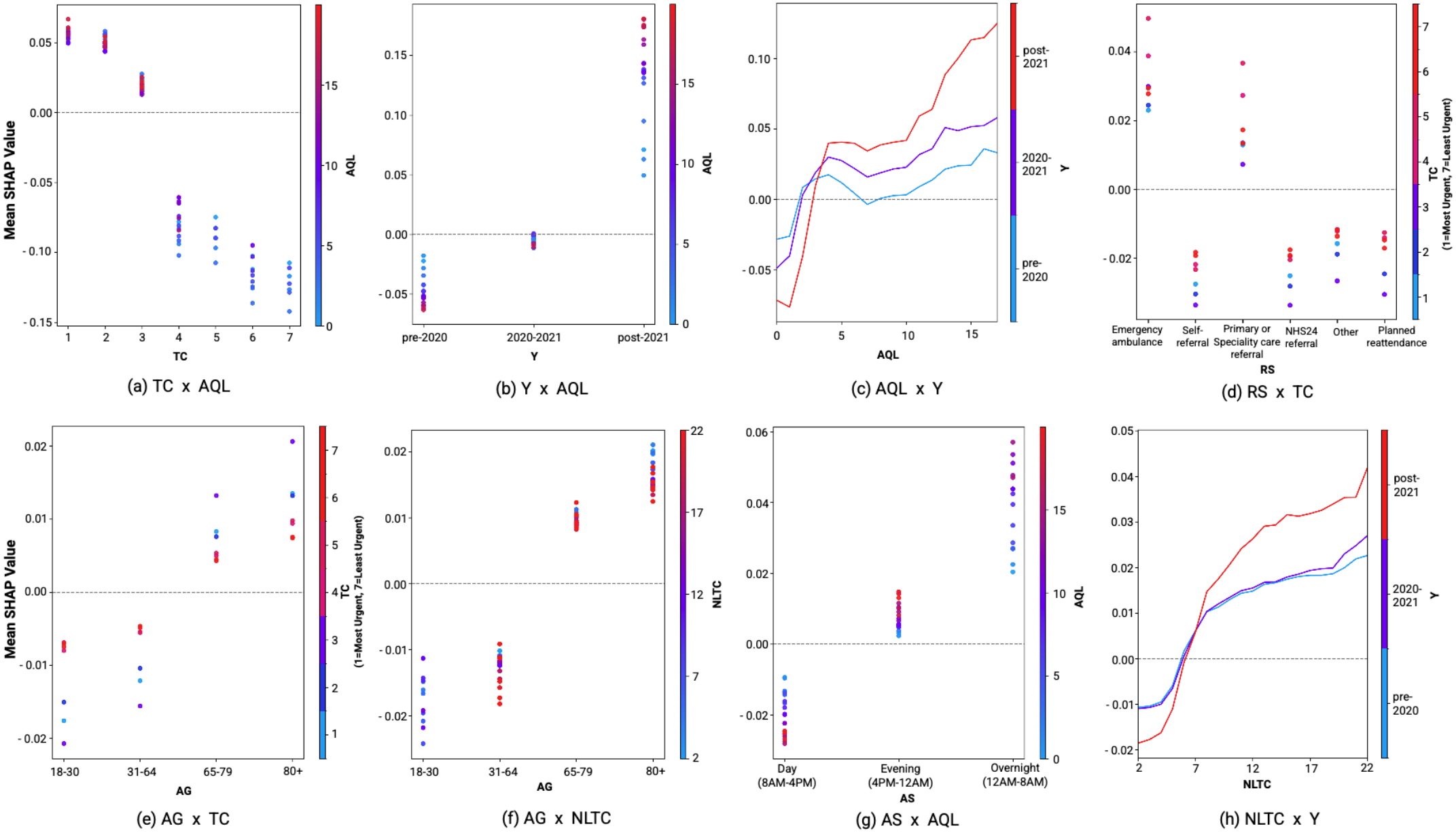
SHAP dependence plots for the four most influential predictors of deviation severity. Each point represents an ED attendance, with colour indicating the value of the interacting feature.

Triage category in Fig. 7(a) shows a strong gradient: highacuity codes (e.g., Code 1: Immediate Resuscitation) were linked to higher deviation, while low-acuity categories (e.g., Code 7: Suitable for Redirection) had increasingly negative effects, relatively stable across queue lengths. Year, in Fig. 7(b), was associated with increasing SHAP contributions from pre-2020 to post-2021. Attendances in the post-2021 period showed markedly higher SHAP values, with a broader range (0.05 to 0.18) compared to 2020-2021 (-0.01 to 0.00) and pre-2020 (-0.07 to -0.01), particularly under high queue pressure. Activity queue length in Fig. 7(c) had a growing influence across all years, with SHAP values turning positive at 2 patients queuing for ED clinical activities (pre-2021 & 2020-2021) and 3 (post-2021). For queues longer than 4 patients, post-2021 attendances showed a stronger contribution to deviation than earlier periods, with an even greater rise observed when queue length reached 12-13 patients.

For referral source in Fig. 7(d), emergency ambulance and GP/specialist referrals were associated with higher deviation, particularly in low-acuity triage groups. To explore age group, two dependence plots were reported. First, with triage category (Fig. 7(e)), older patients in urgent categories ((Code 1-3)) showed higher SHAP values. Second, for condition count (Fig. 7(f)), SHAP values generally increased as the number of LTC increased across age groups. However, for patients aged 80+, deviation severity was more strongly associated with fewer long-term conditions, suggesting that unexpected low complexity in this age group may drive process deviation.

Arrival shift (Fig. 7(g)) revealed that overnight attendances under high queue pressure had the strongest positive deviation impact, whereas daytime visits had lower or negative effects at similar queue levels. When looking at the number of LTCs (Fig. 7(h)), more conditions generally led to more positive SHAP values, especially in the post-2021 period. Across all three year periods, SHAP values turned positive at around 6 MLTC.

## V. Discussion

This study introduced a novel, interpretable, and data-driven framework to quantify deviation at the process-level and explain it through its clinical and operational drivers in ED care for patients with MLTC. The proposed framework integrates multi-perspective conformance checking and explainable machine learning. The multi-perspective conformance checking captures both control-flow misalignments and breaches of the 4-hour ED standard. A new metric named deviation score was introduced through this approach, providing a unified measure of deviation severity. SHAP explainability was applied to identify and interpret the key clinical, sociodemographic, and operational factors associated with higher deviation severity.

### A. Process-Level Validation of MLTC Complexity

The differences observed in Section IV between MLTC and non-MLTC attendances are consistent with well-established clinical and operational challenges in emergency care. MLTC patients were older and presented with higher triage acuity, increasing the likelihood of serious underlying pathology [25]. This greater degree of clinical complexity [10] leads to more urgent patterns of presentation, evident in the higher rates of ambulance arrival, unscheduled referrals, and assignment to higher-acuity triage categories. The higher use of blood tests, bed requests, and inpatient admissions in the MLTC cohort aligns with evidence that multimorbidity naturally requires broader investigation, more time for assessment and treatment, and more frequent specialist involvement [12]. Each added investigation or handover introduces delay, contributing to the longer inter-activity intervals observed in MLTC workflows and consistent with existing studies showing that greater clinical complexity slows progression in the ED settings [20]. The longer total ED durations and higher proportion of MLTC attendances not meeting the 4-hour target reflect the cumulative effect of diagnostic complexity compounded by operational pressures. Our conformance results provide a process-level validation of these differences. Lower conformance rates and higher deviation scores among MLTC attendances indicate more frequent structural and temporal divergences from the optimised pathway. Our metric, the DS, captures clinically plausible and literature-consistent differences between MLTC and non-MLTC cohorts.

### B. Correlation Analysis and Model Reliability

The correlation findings provide an initial view of how clinical, operational, and sociodemographic attributes relate to deviation severity, with the strongest associations aligning with well-established drivers of ED workflow complexity and delay [20], [12], [25]. A moderate correlation was observed between age and number of LTCs, consistent with the expected age-related rise in MLTC burden [10]. VIF analysis confirmed that multicolinearity was not a concern among the predictors included in the final model. However, differences between the correlation analysis and the model-based SHAP explanations are expected, as these methods capture fundamentally different aspects of the data. Spearman’s rho and mutual information quantify only bivariate associations, while the Random Forest model learns multivariate, nonlinear, and interaction effects across the full feature set.

Our findings suggest that the RF regression achieved acceptable performance for modelling deviation severity. As the aim of this study is explanatory rather than predictive, the coefficient of determination provides the most meaningful indication of model adequacy [46]. In complex clinical modelling tasks, the obtained *R*^2^ values are considered significant, supporting the reliability of the SHAP-based interpretations. The MAE values (*<* 0.10 on a normalised scale) indicate that average prediction errors are small in absolute terms, reinforcing the model’s suitability for interpretation even though absolute accuracy is not the primary objective. The sensitivity analysis results indicate that the clustered pattern effectively captured temporal variation, likely reflecting the operational and systemic shifts associated with the COVID-19 pandemic.

### C. Feature-Level Insights into Deviation

SHAP results identified triage acuity, year, activity queue length, and referral source as key drivers of deviation in the MLTC cohort, capturing both clinical urgency and system-level pressures known to increase ED workflow complexity [20], [10]. Age group, arrival shift, and number of LTCs also contributed meaningfully, reflecting known challenges in caring for complex patients. Older adults often require nuanced assessment, and overnight arrivals may be treated by less experienced staff with more distant senior supervision. Whereas all modelled attendances came from the MLTC cohort, those with the highest number of conditions likely needed more care coordination, increasing deviation risk. In contrast, sociodemographic factors like SIMD, sex, and ethnicity had minimal impact, consistent with UK evidence that their influence fades when clinical and operational needs are considered [24].

Both queue length and MLTC showed clear threshold effects, where their contributions to deviation severity shifted from negative to strongly positive. This pattern reflects a non-linear and amplifying impact on process disruption once these factors exceed certain levels. Existing literature supports this observation, showing that ED workflows often remain stable under manageable complexity or demand but become fragile when care demands go beyond system capacity or coordination becomes overly intricate [7], [9], [10], [12], [29], [47]. Our analysis suggests a shift in the system adaptation since COVID, with greater deviation in performance post-pandemic for apparently similar patient burden. This could reflect system changes outside of ED control, where access to hospital beds to allow flow of patients out of ED may be constrained by delayed hospital discharges.

Triage category, year, arrival shift, and number of LTCs represent a consistent directional trend, where higher levels within each domain associated with greater contributions to deviation severity. This likely reflects rising clinical and operational complexity, as urgent cases, post-pandemic constraints, and overnight arrivals introduce challenges that push care beyond routine protocols [47]. Similarly, patients with greater multimorbidity often require tailored management, increasing the likelihood of deviation when system resources are stretched.

Interestingly, patients in moderate-acuity categories (such as urgent (Code 3), standard (Code 4) and not urgent (Code 5)) referred via emergency ambulance or specialist services, as well as those aged 65 and above, showed the highest deviation contributions. These patterns may reflect presentations that initially appear manageable but involve underlying complexity or diagnostic uncertainty [48]. In older adults, subtle symptoms or atypical deterioration can complicate triage and care planning [25]. Notably, among patients aged 80 and over, those with fewer recorded conditions contributed more to deviation than their more MLTC peers. This may reflect appropriate clinical escalation in seemingly lower-risk cases, or conversely reduced prioritisation of these apparently low-risk patients resulting in delays to ED discharge.

### D. Limitations and Future Work

This initial study provides a systematic structural and temporal assessment of ED process deviations based on routinely available operational data, whereas identifying opportunities for methodological refinement and extension. (i) The conformance assessment used a fixed weighting parameter (*α* = 0.5) to balance structural and temporal components when computing DS. This symmetric weighting was intentionally adopted as a baseline, avoiding data-driven optimisation that could compromise interpretability in this initial methodological study. Although equal weighting aligns with practices in related domains [45], its contextual appropriateness in the ED setting has not been empirically calibrated. Future research should explore how different values of *α* affect deviation patterns and their interpretability, potentially informing a more evidence-driven approach to weighting. (ii) Multimorbidity was defined using a simple definition of two or more long-term conditions, which may not fully capture the clinical heterogeneity within the MLTC cohort. Future work should adopt more granular characterisations, such as distinguishing between low and high condition counts or physical-only versus physical–mental health combinations. This would enable a richer examination of how deviation patterns vary across different MLTC profiles.

### E. Significance and Implications

The current study’s findings have important implications towards understanding and addressing the drivers of care pathway deviations for people with MLTC presenting to the ED. In particular, the identified threshold effects highlight critical tipping points where care becomes more fragile and variable. Our findings provide an evidence-based foundation for supporting targeted system adjustments, such as optimising staffing patterns during peak queue times, to reduce deviation risk and improve care consistency. Our proposed framework offers a systematic approach to quantifying how care pathways diverge from expected standards across both process and temporal dimensions, supporting future efforts to evaluate and optimise ED care for people with MLTC. This work forms part of a broader research effort to enhance how urgent care systems respond to the needs of people with MLTC through the application of process-aware analytical methods.

## VI. Conclusion

The present study proposed a process-aware analytical framework to quantify deviation at the process level and explain process deviation in emergency care for people with MLTC. By integrating multi-perspective conformance checking with explainable machine learning, the framework enables both process-level and feature-level insights into deviation patterns. Central to this is the introduction of the deviation score (DS), a new metric that captures structural misalignments and time-based breaches in a unified, interpretable form. Our current findings evidence that attendances with MLTC are characterised by longer and more complex pathways, higher DS, and substantially lower conformance than those without MLTC. Among MLTC patients, deviation severity was primarily driven by triage acuity, year, activity queue length, and referral source. Older age, overnight arrival, and higher multimorbidity burden were also associated with increased deviation severity. Older patients or referral via emergency or specialist routes with moderate-acuity level may deviate more due to hidden complexity in presentations that appear initially stable. Importantly, the uncovered threshold effects in queue length and multimorbidity further reveal critical tipping points beyond which care becomes harder to standardise.

In future work, refining the weighting scheme used in DS and adoption of more nuanced multimorbidity definitions will enable deeper insights into deviation patterns across the MLTC cohort. More broadly, our study highlights the value of combining process- and feature-level perspectives to improve understanding of ED care for people with MLTC.

## Data Availability

The data used in this study were accessed via DataLoch (https://dataloch.org), a secure data repository hosted by NHS Lothian and the University of Edinburgh. Due to patient confidentiality and data governance regulations, the raw clinical data are not publicly available, but researchers may apply for access directly through DataLoch subject to appropriate ethical and governance approvals.

## Acknowledgment

This work was supported by National Institute for Health and Care Research (NIHR) Artificial Intelligence and Multimorbidity: Clustering in Individuals, Space and Clinical Context (AIM-CISC) grant NIHR202639. The views expressed are those of the authors and not necessarily those of the NIHR or the Department of Health and Social Care. This work was also supported by DataLoch (https://dataloch.org/), which is funded by the Data Driven Innovation programme within the Edinburgh and South East Scotland City Region Deal.

## Notes

### Competing Interest Statement

The authors have declared no competing interest.

### Author Declarations

The NHS Lothian DataLoch Access Review Service and the NHS Lothian Caldicott Guardian gave governance approval and waived individual patient consent for this work (DataLoch Project Reference: DL_2022_029), which was conducted under the overarching research ethics approval granted to DataLoch by the South East Scotland Research Ethics Service, as only de-identified routine healthcare data were analysed.

## References

[1] K. Khunti, H. Sathanapally, and P. Mountain, “Multiple long term conditions, multimorbidity, and co-morbidities: we should reconsider the terminology we use,” BMJ, vol. 383, 2023. [Online]. Available: https://www.bmj.com/content/383/bmj.p2327

[2] C. Imison, “Making sense of the evidence: Multiple long-term conditions (multimorbidity),” NIHR Evidence, 03 2021.

[3] A. Gill, K. Kuluski, L. Jaakkimainen, G. Naganathan, R. Upshur, and W. P. Wodchis, ““where do we go from here?” health system frustrations expressed by patients with multimorbidity, their caregivers and family physicians,” Healthcare Policy, vol. 9, pp. 73–89, 04 2014. [Online]. Available: https://www.ncbi.nlm.nih.gov/pmc/articles/PMC4749886/

[4] K. M. Foo, M. Sundram, and H. Legido-Quigley, “Facilitators and barriers of managing patients with multiple chronic conditions in the community: a qualitative study,” BMC Public Health, vol. 20, 02 2020.

[5] J. Valabhji, E. Barron, A. Pratt, N. Hafezparast, R. Dunbar-Rees, E. B. Turner, K. Roberts, J. Mathews, R. Deegan, V. Cornelius, J. Pickles, G. Wainman, C. Bakhai, D. G. Johnston, E. W. Gregg, and K. Khunti, “Prevalence of multiple long-term conditions (multimorbidity) in england: a whole population study of over 60 million people,” Journal of the Royal Society of Medicine, vol. 117, 10 2023. [Online]. Available: https://journals.sagepub.com/doi/10.1177/01410768231206033

[6] G. Simpson, I. Okamoto, L. Morrison, M. Santer, A. Farmer, and H. Dambha-Miller, “Exploring patient, informal carer and care professional perspectives to understand the key clinical and non-clinical care needs of those living with multiple long-term conditions (mltc),” British Journal of General Practice, vol. 73, pp. bjgp23X733 653–bjgp23X733 653, 07 2023. [Online]. Available: https://bjgp.org/content/73/suppl1/bjgp23X733653

[7] L. Dismore, C. Hurst, A. Granic, E. Tullo, M. D. Witham, R. M. Dodds, A. A. Sayer, and S. M. Robinson, “Why are older adults living with the complexity of multiple long-term conditions, frailty and a recent deterioration in health under-served by research? a narrative synthesis review of the literature,” Journal of frailty, sarcopenia and falls, vol. 8, pp. 230–239, 12 2023. [Online]. Available: https://www.ncbi.nlm.nih.gov/pmc/articles/PMC10690133/

[8] K. Moffat and S. W. Mercer, “Challenges of managing people with multimorbidity in today’s healthcare systems,” BMC Family Practice, vol. 16, 2015. [Online]. Available: https://bmcprimcare.biomedcentral.com/articles/10.1186/s12875-015-0344-4

[9] H. Hassankhani, A. Soheili, S. S. Vahdati, F. Amin Mozaffari, L. A. Wolf, and T. Wiseman, ““me first, others later” a focused ethnography of ongoing cultural features of waiting in an iranian emergency department,” International Emergency Nursing, vol. 47, p. 100804, 11 2019.

[10] C. Hudon, K. Aubrey-Bassler, M.-C. Chouinard, S. Doucet, M.-F. Dubois, M. Karam, A. Luke, G. Moullec, P. Pluye, A. Tzenov, S. Ouadfel, M. Lambert, E. Angrignon-Girouard, C. Schwarz, D. Howse, K. K. MacLeod, A. Gaudreau, and V. Sabourin, “Better understanding care transitions of adults with complex health and social care needs: a study protocol,” BMC Health Services Research, vol. 22, 02 2022. [Online]. Available: https://bmchealthservres.biomedcentral.com/articles/10.1186/s12913-022-07588-0

[11] J. F. Mize, “The roundabout way to disaster: Recognizing and responding to normalization of deviance,” Process Safety Progress, vol. 38, p. e12014. 11 2018.

[12] M. C. Blayney, M. J. Reed, J. A. Masterson, A. Anand, M. M. Bouamrane, J. Fleuriot, S. Luz, M. J. Lyall, S. Mercer, N. L. Mills, S. D. Shenkin, T. S. Walsh, S. H. Wild, H. Wu, S. McLachlan, B. Guthrie, and N. I. Lone, “Multimorbidity and adverse outcomes following emergency department attendance: population based cohort study,” BMJ Medicine, vol. 3, p. e000731. 08 2024. [Online]. Available: https://bmjmedicine.bmj.com/content/bmjmed/3/1/e000731.full.pdf

[13] C. J. Cohen-Stavi, S. Giveon, C. Key, T. Molcho, R. Balicer, and E. Shadmi, “Guideline deviation and its association with specific chronic diseases among patients with multimorbidity: a cross-sectional cohort study in a care management setting,” BMJ Open, vol. 11, p. e040961. 01 2021.

[14] K. J. O Connell, K. N. Shaw, R. M. Ruddy, P. V. Mahajan, R. Lichenstein, C. S. Olsen, T. Funai, S. Blumberg, and J. M. Chamberlain, “Incident reporting to improve patient safety,” Pediatric Emergency Care, vol. 34, pp. 237–242, 04 2018.

[15] S. Jones, C. Moulton, S. Swift, P. Molyneux, S. Black, N. Mason, R. Oakley, and C. Mann, “Association between delays to patient admission from the emergency department and all-cause 30-day mortality,” Emergency Medicine Journal, vol. 39, pp. 168–173, 01 2022. [Online]. Available: https://emj.bmj.com/content/early/2022/01/03/emermed-2021-211572

[16] S. Yang, A. Sarcevic, R. A. Farneth, S. Chen, O. Z. Ahmed, I. Marsic, and R. S. Burd, “An approach to automatic process deviation detection in a time-critical clinical process,” Journal of Biomedical Informatics, vol. 85, pp. 155–167, 09 2018.

[17] N. Doutcheva, M. N. Shah, A. Borkenhagen, M. K. Finta, J. Duckles, C. R. Sellers, S. Seshadri, D. Lampo, and N. E. Werner, “Process variances in older adults’ care transitions from emergency department to home: Process breakdown versus process resiliency,” Proceedings of the Human Factors and Ergonomics Society Annual Meeting, vol. 61, pp. 565–566, 09 2017.

[18] T. G. Erdogan and A. K. Tarhan, “Multi-perspective process mining for emergency process,” Health Informatics Journal, vol. 28, p. 146045822210771, 01 2022.

[19] A. Celda-Moret, G. Ibanez-Sanchez, J. Garijo, M. Pop-Llut, M. Faus-Lluquet, and C. Fernandez-Llatas, “Fast track design using process mining: Does it improve saturation and times in emergency departments?” Applied Sciences, vol. 15, no. 13, 2025. [Online]. Available: https://www.mdpi.com/2076-3417/15/13/7367

[20] M. Cho, M. Song, J. Park, S.-R. Yeom, I.-J. Wang, and B.-K. Choi, “Process mining-supported emergency room process performance indicators,” International Journal of Environmental Research and Public Health, vol. 17, p. 6290, 08 2020.

[21] H. Wang, N. Sambamoorthi, D. Sandlin, and U. Sambamoorthi, “Interpretable machine learning models for prolonged emergency department wait time prediction,” BMC Health Services Research, vol. 25, 03 2025.

[22] E. Arnaud, P. A. Moreno-Sanchez, M. Elbattah, C. Ammirati, M. van Gils, G. Dequen, and D. A. Ghazali, “Development and clinical interpretation of an explainable AI model for predicting patient pathways in the emergency department: A retrospective study,” Applied Sciences, vol. 15, no. 15, 2025. [Online]. Available: https://www.mdpi.com/2076-3417/15/15/8449

[23] W. A. Sulaiman, C. Stylianides, A. Nikolaou, Z. Antoniou, I. Constantinou, L. Palazis, A. Vavlitou, T. Kyprianou, E. Kyriacou, A. Kakas, M. S. Pattichis, A. S. Panayides, and C. S. Pattichis, “Leveraging machine learning and rule extraction for enhanced transparency in emergency department length of stay prediction,” Frontiers in Digital Health, vol. Volume 6 - 2024, 2025. [Online]. Available: https://www.frontiersin.org/journals/digital-health/articles/10.3389/fdgth.2024.1498939

[24] C. McParland, M. A. Cooper, D. J. Lowe, B. Stanley, and B. Johnston, “Multimorbidity, disease count, mortality and emergency care use in persons attending the emergency department: a cross-sectional data-linkage study,” Journal of Multimorbidity and Comorbidity, vol. 12, p. 26335565221147417, 2022. [Online]. Available: 10.1177/26335565221147417

[25] Y. Yordanov, A. Beauvais, and P.-C. Thiébaud, “Multimorbidity in emergency departments: urgent need for integrated care,” BMJ Medicine, vol. 3, p. e000989. 08 2024.

[26] A. Wartelle, F. Mourad-Chehade, F. Yalaoui, H. Questiaux, T. Monneret, G. Soliveau, J. Chrusciel, A. Duclos, D. Laplanche, and S. Sanchez, “Multimorbidity clustering of the emergency department patient flow: Impact analysis of new unscheduled care clinics,” PLOS ONE, vol. 17, p. e0262914. 01 2022.

[27] A. Lim, C. Benjasirisan, X. Liu, O. Ogungbe, C. D. Himmelfarb, P. Davidson, and B. Koirala, “Social determinants of health and emergency department visits among older adults with multimorbidity: insight from 2010 to 2018 national health interview survey,” BMC public health, vol. 24, 04 2024.

[28] C. MacRae, H. W. Fisken, E. Lawrence, T. Connor, J. Pearce, A. Marshall, A. Lawson, C. Dibben, S. W. Mercer, and B. Guthrie, “Household and area determinants of emergency department attendance and hospitalisation in people with multimorbidity: a systematic review,” BMJ Open, vol. 12, p. e063441. 10 2022. [Online]. Available: https://bmjopen.bmj.com/content/12/10/e063441

[29] N. L. Howe, E.-R. Blackburn, A. Sheppard, S. Pretorius, J. Suklan, S. Bellass, R. Cooper, S. Gallier, E. Sapey, A. A. P. Sayer, and M. Witham, “Clinical decision-making and care pathways for people with multiple long-term conditions admitted to hospital: a scoping review,” BMJ Open, vol. 15, no. 8, 2025. [Online]. Available: https://bmjopen.bmj.com/content/15/8/e100270

[30] W. van der Aalst, Data Science in Action. Berlin, Heidelberg: Springer Berlin Heidelberg, 2016, pp. 3–23. [Online]. Available: 10.1007/978-3-662-49851-41

[31] W. Yang and Q. Su, “Process mining for clinical pathway: Literature review and future directions,” in 2014 11th International Conference on Service Systems and Service Management (ICSSSM), 2014, pp. 1–5.

[32] H. Al-Badarneh and A. Arif, “A systematic review of process mining in healthcare,” in 2025 International Conference on New Trends in Computing Sciences (ICTCS), 2025, pp. 485–492.

[33] D. Duma and R. Aringhieri, “Mining the patient flow through an emergency department to deal with overcrowding,” Springer Proceedings in Mathematics & Statistics, vol. 210, pp. 49–59, 2017.

[34] E. Rojas, A. Cifuentes, A. Burattin, J. Munoz-Gama, M. Sepúlveda, and D. Capurro, “Performance analysis of emergency room episodes through process mining,” International Journal of Environmental Research and Public Health, vol. 16, p. 1274, 04 2019.

[35] E. Oliart, E. Rojas, and D. Capurro, “Are we ready for conformance checking in healthcare? measuring adherence to clinical guidelines: A scoping systematic literature review,” Journal of Biomedical Informatics, vol. 130, p. 104076, 2022. [Online]. Available: https://www.sciencedirect.com/science/article/pii/S1532046422000922

[36] E. Asare, L. Wang, and X. Fang, “Conformance checking: Workflow of hospitals and workflow of open-source emrs,” IEEE Access, vol. 8, pp. 139546–139566, 2020. [Online]. Available: https://ieeexplore.ieee.org/abstract/document/9149860

[37] L. Nannini, A. C. Bolos, M. Lama, and S. Barro, “Explainable ai in process mining: A systematic literature review,” 01 2025. [Online]. Available: https://ssrn.com/abstract=5162771

[38] J. M. Juarez, Explainable Artificial Intelligence and Process Mining Applications for Healthcare. Springer Nature, 2023.

[39] N. Mehdiyev, M. Majlatow, and P. Fettke, “Interpretable and explainable machine learning methods for predictive process monitoring: A systematic literature review,” 12 2023. [Online]. Available: https://arxiv.org/pdf/2312.17584

[40] V. Kuan, S. Denaxas, A. Gonzalez-Izquierdo, K. Direk, O. Bhatti, S. Husain, S. Sutaria, M. Hingorani, D. Nitsch, C. A. Parisinos, R. T. Lumbers, R. Mathur, R. Sofat, J. P. Casas, I. C. K. Wong, H. Hemingway, and A. D. Hingorani, “A chronological map of 308 physical and mental health conditions from 4 million individuals in the english national health service,” The Lancet Digital Health, vol. 1, pp. e63–e77, 06 2019.

[41] S. Government, “Simd (scottish index of multiple deprivation),” Scottish Index of Multiple Deprivation, 2020. [Online]. Available: https://simd.scot/#/simd2020/BTTTFTT/9/-4.0000/55.9000/

[42] T. Murata, “Petri nets: Properties, analysis and applications,” Proceedings of the IEEE, vol. 77, pp. 541–580, 04 1989.

[43] A. Berti, S. van Zelst, and D. Schuster, “Pm4py: A process mining library for python,” Software Impacts, vol. 17, pp. 100556–100556, 09 2023.

[44] W. van der Aalst, A. Adriansyah, and B. van Dongen, “Replaying history on process models for conformance checking and performance analysis,” WIREs Data Mining and Knowledge Discovery, vol. 2, pp. 182–192, 01 2012.

[45] O. Dogan and H. Avvad, “Fuzzy clustering based on activity sequence and cycle time in process mining,” Axioms, vol. 14, no. 5, 2025. [Online]. Available: https://www.mdpi.com/2075-1680/14/5/351

[46] D. Chicco, M. J. Warrens, and G. Jurman, “The coefficient of determination r-squared is more informative than smape, mae, mape, mse and rmse in regression analysis evaluation,” PeerJ Computer Science, vol. 7, p. e623, 2021. [Online]. Available: https://peerj.com/articles/cs-623/

[47] T. Whiteside, E. Kane, B. Aljohani, M. Alsamman, and A. Pourmand, “Redesigning emergency department operations amidst a viral pandemic,” The American Journal of Emergency Medicine, vol. 38, pp. 1448–1453, 07 2020.

[48] M. Puig-Campmany and J. Ris-Romeu, “Older adults and low-severity emergencies: oversue of hospital emergency services?” Emergencias : revista de la Sociedad Espanola de Medicina de Emergencias, vol. 37, pp. 244–246, 06 2025. [Online]. Available: https://pubmed.ncbi.nlm.nih.gov/40887909/

